# Seroprevalence of SARS-CoV-2 IgG in healthcare workers and other staff at North Bristol NHS Trust: a sociodemographic analysis

**DOI:** 10.1101/2020.11.12.20230458

**Authors:** Christopher R. Jones, Fergus W. Hamilton, Ameeka Thompson, Tim T. Morris, Ed Moran

## Abstract

**Background:** Healthcare workers (HCWs) are at increased risk of infection with Severe Acute Respiratory Syndrome Coronavirus 2 (SARS-CoV-2). There are limited data exploring the relative impact of geographical and socioeconomic factors on risk of SARS-CoV-2 infection among HCWs.

**Aim:** To estimate and explore SARS-CoV-2 IgG antibody seroprevalence in HCWs and support staff at a hospital in South West England.

**Methods:** We conducted a nested cross-sectional study from May-July 2020. Inverse probability weighted regression was used to estimate seroprevalence of SARS-CoV-2 and associations with demographic and socioeconomic risk factors that were robust to selection into testing.

**Findings:** Attendance for testing varied by demographic and socioeconomic factors. The overall rate of SARS-CoV-2 IgG seroprevalence among tested staff was 9.3% (638/6858). The highest seroprevalence was found in wards associated with SARS-CoV-2 outbreaks and among junior staff with patient-facing roles. Black, Asian and Minority Ethnic (BAME) staff had increased odds of SARS-CoV-2 seroprevalence (adjusted OR: 1.99, 95%CI: 1.69, 2.34; p<0.001) relative to White staff, except for those categorised as Medical/Dental. We found a significant association between neighbourhood deprivation and seroprevalence (p<0.01). Seroprevalence ranged from 12% in staff residing in areas with the greatest relative deprivation to 8.4% in the least deprived.

**Conclusion:** Transmission between staff groups is evident within the healthcare setting. BAME individuals were at increased risk of infection with SARS-CoV-2. Work role, area of residence, and neighbourhood deprivation all contribute to SARS-CoV-2 infection risk. As hospitals introduce routine staff SARS-CoV-2 testing they should consider differential uptake of testing among staff groups.

## Introduction

Severe Acute Respiratory Syndrome Coronavirus 2 (SARS-CoV-2) causes coronavirus disease 2019 (COVID-19). It was first identified in Wuhan, China in December 2019 and rapidly spread around the world^1^. To date, over 45 million cases and 1.2 million deaths have been recorded globally^2^. All areas of the United Kingdom (UK) have experienced significant healthcare-related SARS-CoV-2 infection in both hospitals^3^ and care homes^4^. Hospital Trusts quickly introduced universal personal protective equipment (PPE) for staff and other infection control measures, which were associated with a reduced risk of hospital-acquired infections^5-7^. Despite this, healthcare workers (HCWs) have been found to have an increased risk of SARS-CoV-2 infection relative to the general population^7^. It is important to understand which staff groups have the highest risk of SARS-CoV-2 infection so that measures can be taken to protect those at high occupational risk.

Variation in the local burden of disease in the UK was previously shown to be strongly associated with area of residence and neighbourhood deprivation^8^. Between 1^st^ March 2020 and 30^th^ June 2020, the age-standardised mortality rate of deaths involving COVID-19 in the most deprived areas of England was more than double that in the least deprived areas (139.6 deaths/100 000 population versus 63.4 deaths/100 000 population, respectively)^9^. Published studies on SARS-CoV-2 infection in HCWs have highlighted differential rates of seroprevalence and posited that this is linked to exposure within the workplace. Thus far, there are limited data exploring the relative impact of geographical and socioeconomic factors on risk of SARS-CoV-2 infection among HCWs.

The aim of this study was to estimate SARS-CoV-2 IgG antibody seroprevalence amongst HCWs and support staff at a large teaching hospital in South West England and explore differences in rates according to demographic and socioeconomic characteristics. Many published studies on SARS-CoV-2 have been based on highly selected samples and are therefore at risk of selection bias induced by non-random testing patterns amongst volunteers^10^. Here, we use a nested study design to obtain seroprevalence results that are robust to selection bias.

## Materials and Methods

### Study design and setting

This was a cross-sectional SARS-CoV-2 IgG antibody seroprevalence study of HCWs and support staff at North Bristol NHS Trust. The Trust provides inpatient care in a 900-bed hospital that covers a population of approximately 500 000 people in Bristol, South Gloucestershire, and North Somerset. The Trust also provides tertiary services to a population of approximately 1.1 million people in the wider South West England region. The Trust employed 9317 permanent and fixed term staff during the period that this study was conducted. A further 2085 bank staff and 852 staff members who left employment in the 6 months preceding this study were included (total =12 254).

### Data collection

In May 2020, all employees were invited by Trust-wide email communications and posters to self-register for a voluntary SARS-CoV-2 antibody test. The results of these tests were cross-referenced with selected information extracted from the employee records of all members of staff who worked at North Bristol NHS Trust between January 2020 and June 2020. This included: age; self-identified ethnicity; gender; role; ward/department; division; presence or absence of a SARS-CoV-2 serology test; serology result; and postcode. All data were pseudonymised and stored in a password protected folder on the NHS servers with restricted access. An employee number was used to link data sets and ensure no duplication of permanent and bank staff members.

### SARS-CoV-2 antibody assay

The serological status of HCWs and support staff was determined using one of two platforms: 1) the Abbott™ SARS-CoV-2 IgG chemiluminescent microparticle assay on an Architect™ system (Abbott Laboratories); or 2) the Roche™ Elecsys® Anti-SARS-CoV-2 (IgG/IgM) electrochemiluminescent immunoassay on a Cobas™ analyser (Roche Diagnostics).

### Statistical analysis

Data were subject to descriptive analysis with n (%) or median (IQR) being presented, as appropriate. Data were first analysed according to testing status to determine selection into the testing sample across measured characteristics including age group, ethnicity, gender, staff role, and residential neighbourhood deprivation. To preserve anonymity, office-based administrative staff groups were re-labelled using generic descriptive terms. The SARS-CoV-2 serology results were then compared across groups using chi-square test of independence or Wilcoxon rank-sum test. We used inverse probability weighting (IPW) to standardise the tested sample to the full North Bristol NHS Trust workforce. By weighting estimates amongst those tested by their probability of being tested, IPW can recover unbiased estimates of associations. Weights were constructed by estimating the likelihood of individuals being selected into the sample from the super-sample based upon measured characteristics^11^. We used weighted regression to estimate associations between risk factors and SARS-CoV-2 seroprevalence.

To investigate spatial variation in testing uptake and seroprevalence, we aggregated data on place of residence to Middle Layer Super Output Areas (MSOAs). MSOAs are geospatial units with an average population of 7200 that are designed to be socially homogeneous. Staff postcodes were matched to MSOAs using the published 2019 boundaries. We used Index of Multiple Deprivation (IMD) as a proxy for socioeconomic position, with IMD deciles matched to MSOAs where 1 is the most deprived decile and 10 the least deprived. The IMD is a composite measure of relative deprivation within a defined small geographic area and covers seven domains – income; employment; education, skills and training; health and disability; crime; barriers to housing and services; and living environment^12^. We extracted data from the 2019 report^13^ compiled by the Office for National Statistics for use in this study. Maps of the wider Bristol and South Gloucestershire area were generated using the R packages “sf” and “ggmaps”. The following data were presented – staff testing by MSOA; staff SARS-CoV-2 seroprevalence by MSOA; PHE data^14^ on rate/100 000 population at MSOA level; and IMD decile by MSOA. All analyses were performed using R (Version 4.0.0).

### Ethics

Ethical approval for this study was granted by the North West - Greater Manchester West Research Ethics Committee (20/NW/0354). Data were processed under the COVID-19 Control of Patient Information (COPI) arrangements (see https://www.gov.uk/government/publications/coronavirus-covid-19-notification-of-data-controllers-to-share-information). All procedures were conducted in line with the Declaration of Helsinki.

## Results

Of the 12 254 North Bristol NHS Trust HCWs and support staff registered during the study period, a total of 6861 (56%) underwent SARS-CoV-2 antibody testing (table I). Three cases were excluded due to incomplete data (tested but no result available) and 6858 were included in the final analysis.

**Table I:** Sociodemographic characteristics of HCWs and support staff attending for SARS-CoV-2 serological testing. One staff group was removed (not tested n=1; tested n=2) to preserve anonymity. Abbreviations: BAME – Black, Asian and Minority Ethnic; IMD – Indices of Multiple Deprivation.

| Characteristic | Not Tested, N = 5393<sup>1</sup> |  | Tested, N = 6858<sup>1</sup> |  | p-value<sup>2</sup> |
| --- | --- | --- | --- | --- | --- |
| Age | n | % | n | % | <0.001 |
| <=20 Years | 142 | 55% | 114 | 45% |  |
| 21-30 | 1540 | 47% | 1757 | 53% |  |
| 31-40 | 1481 | 48% | 1624 | 52% |  |
| 41-50 | 917 | 37% | 1536 | 63% |  |
| 51-60 | 876 | 38% | 1408 | 62% |  |
| 61-70 | 400 | 50% | 402 | 50% |  |
| >=71 Years | 37 | 69% | 17 | 31% |  |
| Gender |  |  |  |  | <0.001 |
| Female | 3839 | 42% | 5338 | 58% |  |
| Male | 1554 | 51% | 1520 | 49% |  |
| Ethnicity |  |  |  |  | <0.001 |
| BAME | 1015 | 48% | 1095 | 52% |  |
| Undisclosed | 289 | 61% | 185 | 39% |  |
| White | 4089 | 42% | 5578 | 58% |  |
| Staff group |  |  |  |  | <0.001 |
| Additional Scientific & Technical | 123 | 37% | 211 | 63% |  |
| Additional Clinical Services | 1363 | 49% | 1420 | 51% |  |
| Administrative & Clerical | 928 | 43% | 1233 | 57% |  |
| Allied Health Professionals | 183 | 31% | 413 | 69% |  |
| Estates & Ancillary | 641 | 55% | 516 | 45% |  |
| Healthcare Scientists | 178 | 42% | 245 | 58% |  |
| Medical & Dental | 892 | 51% | 856 | 49% |  |
| Nursing & Midwifery | 1084 | 36% | 1962 | 64% |  |
| Frontline |  |  |  |  | 0.11 |
| Non-patient facing | 1780 | 45% | 2167 | 55% |  |
| Frontline | 3613 | 44% | 4691 | 56% |  |
| Work schedule |  |  |  |  | <0.001 |
| Bank | 2029 | 81% | 472 | 19% |  |
| Fixed Term Temporary | 546 | 42% | 740 | 58% |  |
| Permanent | 2818 | 33% | 5646 | 67% |  |
| IMD Decile |  |  |  |  | 0.001 |
| 1 (most deprived) | 362 | 49% | 375 | 51% |  |
| 2 | 539 | 45% | 663 | 55% |  |
| 3 | 381 | 44% | 480 | 56% |  |
| 4 | 521 | 46% | 617 | 54% |  |
| 5 | 506 | 45% | 628 | 55% |  |
| 6 | 361 | 43% | 488 | 57% |  |
| 7 | 546 | 42% | 746 | 58% |  |
| 8 | 558 | 44% | 717 | 56% |  |
| 9 | 551 | 44% | 694 | 56% |  |
| 10 (least deprived) | 761 | 40% | 1161 | 60% |  |
| <sup>1</sup>Statistics presented: n (%) |  |  |  |  |  |
| <sup>2</sup>Statistical tests performed: chi-square test of independence; Wilcoxon rank-sum test |  |  |  |  |  |

### Staff uptake of voluntary testing

Volunteering for testing varied by demographic characteristics. Older age groups were more likely to present for testing, with those aged 51-60 (63%) and 61-70 (62%) the most likely; females (58%) were more likely to present than males (49%); White individuals (58%) were more likely to present than BAME (52%); and permanent staff (67%) were more likely to present than bank staff (19%) (all p<0.001). Attendance for testing ranged from 51% in the most deprived decile to 60% in the least deprived (p=0.001 for trend). Testing was similar across frontline and non-patient facing roles (p=0.11). Results for the first stage of the weighted regression analysis with parameter estimates for testing can be found in supplementary table I.

### SARS-CoV-2 IgG seroprevalence in HCWs and support staff

The overall rate of SARS-CoV-2 seroprevalence among tested HCWs and support staff was 9.3% (638/6858) (Table II). Black, Asian and Minority Ethnic individuals were more likely to be seropositive relative to White (14.6% versus 8.2%, respectively; p<0.001). Seroprevalence was similar between females and males (9.3% versus 9.2%, respectively; p=0.9). Seroprevalence generally decreased with age, being highest in those aged ≤20y (12.3%) and lowest in those aged ≥71y (5.9%) (p for trend <0.001). Seroprevalence was highest in the Medicine Division (18.3%), Clinical Governance Division (15.7%) and Bank Staff (14.2%). Seroprevalence ranged from 12.0% in the most deprived decile to 8.4% in the least deprived, with a significant association (p<0.01) between deprivation and seroprevalence. Weighted results were broadly similar, though they suggested that seroprevalence was underestimated in certain staff groups due to non-random differences in presentation for testing. For example, the weighted results estimated seroprevalence amongst BAME staff to be 15.7% rather than 14.6%. Weighted data suggested that seroprevalence was overestimated in the clinical governance division (15.7% unweighted versus 6.4% weighted).

**Table II:**
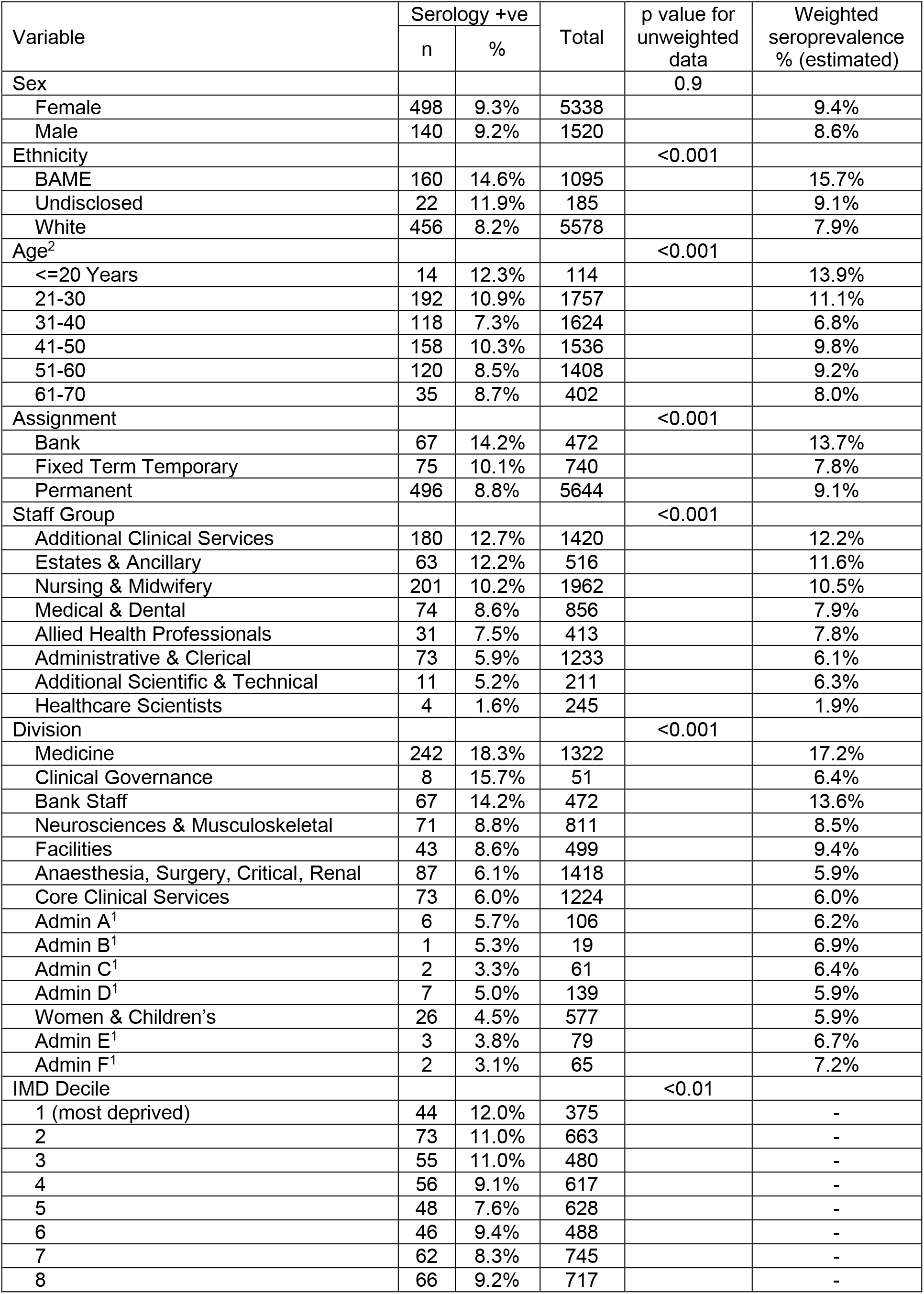
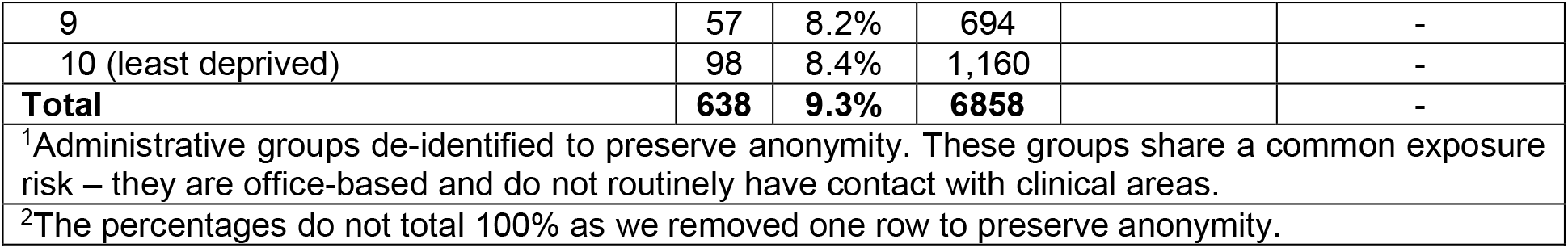
SARS-CoV-2 IgG antibody status of HCWs and support staff according to sociodemographic characteristics. Both unweighted and inverse probability weighted data are presented. The p values were calculated using unweighted data. Abbreviations: +ve – positive; % - proportion; BAME – Black, Asian and Minority Ethnic; IMD – Indices of Multiple Deprivation.

| Variable | Serology +ve |  | Total | p value for unweighted data | Weighted seroprevalence % (estimated) |
| --- | --- | --- | --- | --- | --- |
|  | n | % |  |  |  |
| Sex |  |  |  | 0.9 |  |
| Female | 498 | 9.3% | 5338 |  | 9.4% |
| Male | 140 | 9.2% | 1520 |  | 8.6% |
| Ethnicity |  |  |  | <0.001 |  |
| BAME | 160 | 14.6% | 1095 |  | 15.7% |
| Undisclosed | 22 | 11.9% | 185 |  | 9.1% |
| White | 456 | 8.2% | 5578 |  | 7.9% |
| Age<sup>2</sup> |  |  |  | <0.001 |  |
| <=20 Years | 14 | 12.3% | 114 |  | 13.9% |
| 21-30 | 192 | 10.9% | 1757 |  | 11.1% |
| 31-40 | 118 | 7.3% | 1624 |  | 6.8% |
| 41-50 | 158 | 10.3% | 1536 |  | 9.8% |
| 51-60 | 120 | 8.5% | 1408 |  | 9.2% |
| 61-70 | 35 | 8.7% | 402 |  | 8.0% |
| Assignment |  |  |  | <0.001 |  |
| Bank | 67 | 14.2% | 472 |  | 13.7% |
| Fixed Term Temporary | 75 | 10.1% | 740 |  | 7.8% |
| Permanent | 496 | 8.8% | 5644 |  | 9.1% |
| Staff Group |  |  |  | <0.001 |  |
| Additional Clinical Services | 180 | 12.7% | 1420 |  | 12.2% |
| Estates & Ancillary | 63 | 12.2% | 516 |  | 11.6% |
| Nursing & Midwifery | 201 | 10.2% | 1962 |  | 10.5% |
| Medical & Dental | 74 | 8.6% | 856 |  | 7.9% |
| Allied Health Professionals | 31 | 7.5% | 413 |  | 7.8% |
| Administrative & Clerical | 73 | 5.9% | 1233 |  | 6.1% |
| Additional Scientific & Technical | 11 | 5.2% | 211 |  | 6.3% |
| Healthcare Scientists | 4 | 1.6% | 245 |  | 1.9% |
| Division |  |  |  | <0.001 |  |
| Medicine | 242 | 18.3% | 1322 |  | 17.2% |
| Clinical Governance | 8 | 15.7% | 51 |  | 6.4% |
| Bank Staff | 67 | 14.2% | 472 |  | 13.6% |
| Neurosciences & Musculoskeletal | 71 | 8.8% | 811 |  | 8.5% |
| Facilities | 43 | 8.6% | 499 |  | 9.4% |
| Anaesthesia, Surgery, Critical, Renal | 87 | 6.1% | 1418 |  | 5.9% |
| Core Clinical Services | 73 | 6.0% | 1224 |  | 6.0% |
| Admin A<sup>1</sup> | 6 | 5.7% | 106 |  | 6.2% |
| Admin B<sup>1</sup> | 1 | 5.3% | 19 |  | 6.9% |
| Admin C<sup>1</sup> | 2 | 3.3% | 61 |  | 6.4% |
| Admin D<sup>1</sup> | 7 | 5.0% | 139 |  | 5.9% |
| Women & Children's | 26 | 4.5% | 577 |  | 5.9% |
| Admin E<sup>1</sup> | 3 | 3.8% | 79 |  | 6.7% |
| Admin F<sup>1</sup> | 2 | 3.1% | 65 |  | 7.2% |
| IMD Decile |  |  |  | <0.01 |  |
| 1 (most deprived) | 44 | 12.0% | 375 |  | - |
| 2 | 73 | 11.0% | 663 |  | - |
| 3 | 55 | 11.0% | 480 |  | - |
| 4 | 56 | 9.1% | 617 |  | - |
| 5 | 48 | 7.6% | 628 |  | - |
| 6 | 46 | 9.4% | 488 |  | - |
| 7 | 62 | 8.3% | 745 |  | - |
| 8 | 66 | 9.2% | 717 |  | - |
| 9 | 57 | 8.2% | 694 |  | - |
| 10 (least deprived) | 98 | 8.4% | 1,160 |  | - |
| **Total** | **638** | **9.3%** | **6858** |  | - |
| <sup>1</sup>Administrative groups de-identified to preserve anonymity. These groups share a common exposure risk – they are office-based and do not routinely have contact with clinical areas. |  |  |  |  |  |
| <sup>2</sup>The percentages do not total 100% as we removed one row to preserve anonymity. |  |  |  |  |  |

Given the higher seroprevalence in BAME individuals, we hypothesised that this may be related to profession and explored this with subgroup analysis (Figure 1). Seroprevalence was higher in BAME than White individuals across all staff groups, except for Medical/Dental, where the trend was reversed (4.4% BAME versus 9.6% White). Within the Medical/Dental group, the proportion of BAME individuals at early career stages was smaller (17.1% versus 25.6%) relative to White individuals (supplementary table II). The median IMD decile for BAME staff was 4 (IQR: 2, 7) and for white staff was 7 (IQR: 4, 9) (data not shown). When restricting to medical and dental staff only, the median IMD decile for BAME staff (8; IQR: 4, 9) and for White staff (8; IQR: 6, 9) were similar (data not shown).

**Figure 1:**
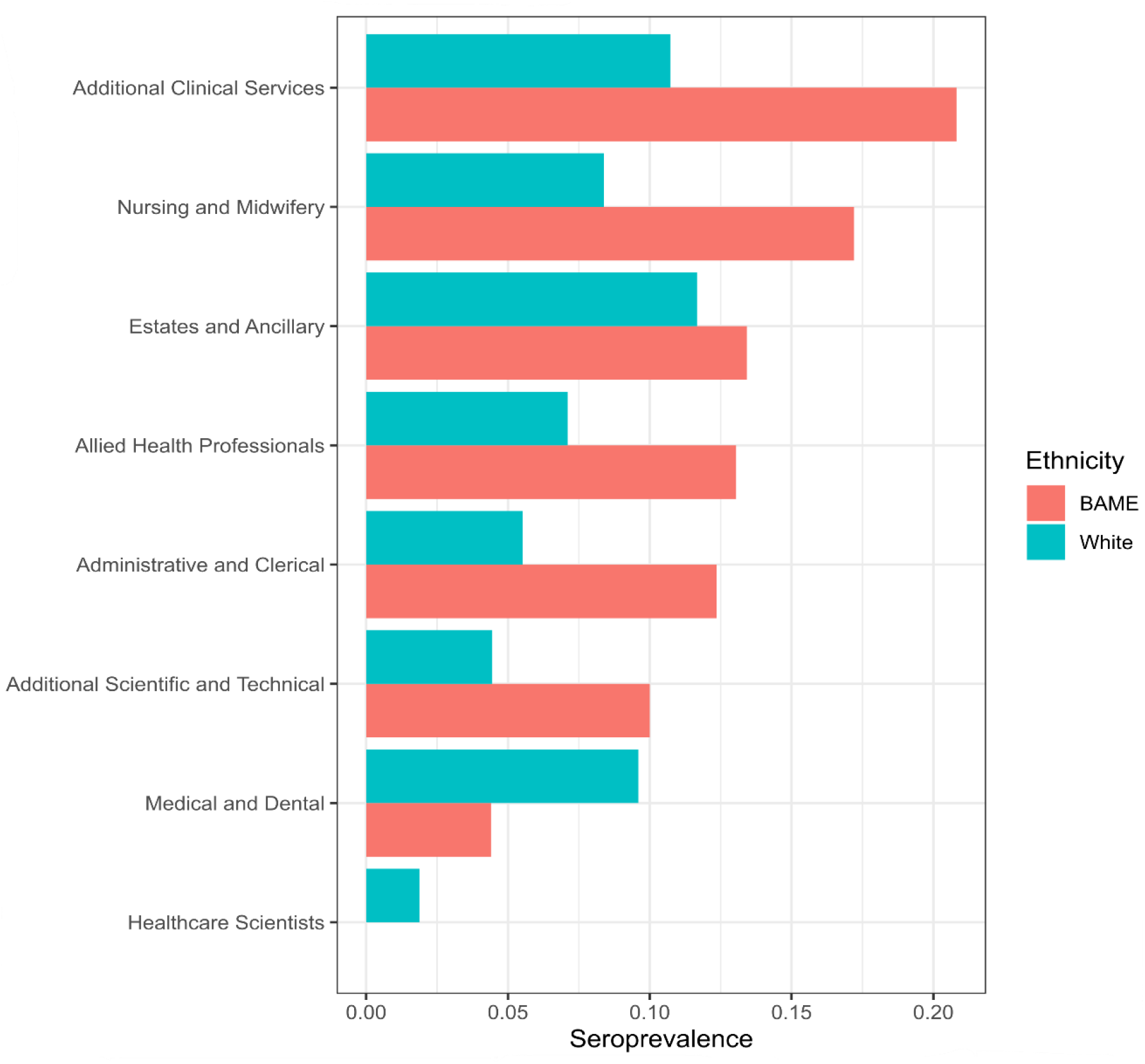
SARS-CoV-2 IgG antibody seroprevalence by staff group and ethnicity. The graph was restricted to HCWs and support staff who disclosed self-identified ethnicity. Abbreviation: BAME – Black, Asian and Minority Ethnic.

We evaluated seroprevalence by inpatient wards (supplementary table III). We found low seroprevalence in staff from the intensive care unit (2.5%). We found 13.6% (respiratory ward, including non-invasive ventilation bays) and 20.9% (elderly care) staff seroprevalence on the two designated COVID-19 inpatient wards. The seroprevalence rate in staff working on the acute medical unit was 16.2%. We found high seroprevalence in staff working in wards that experienced outbreaks – 50% on an elderly care step-down ward and 52.4% on a cardiology ward. The highest rate was found in a satellite dialysis unit (58.8%).

### Geospatial variation in SARS-CoV-2 antibody testing uptake and seroprevalence

We reviewed data on infection rates published by Public Health England (PHE) to contextualise our geographical data ^14^. This revealed differential rates of SARS-CoV-2 infection/100 000 population across the wider Bristol area (Figure 2A-C). The IMD decile according to MSOA is presented in figure 2D. Our data demonstrated variance in staff seroprevalence across areas of residence (Figure 2E). Staff uptake for testing was not affected by area of residence and was consistent across the city (Figure 2F). The highest levels of SARS-CoV-2 staff seroprevalence were in Bath and North East Somerset 020 (30%, n = 16), Bristol 055 (29.4%, n = 76), and North Somerset 009 (21.4%, n = 24).

**Figure 2:**
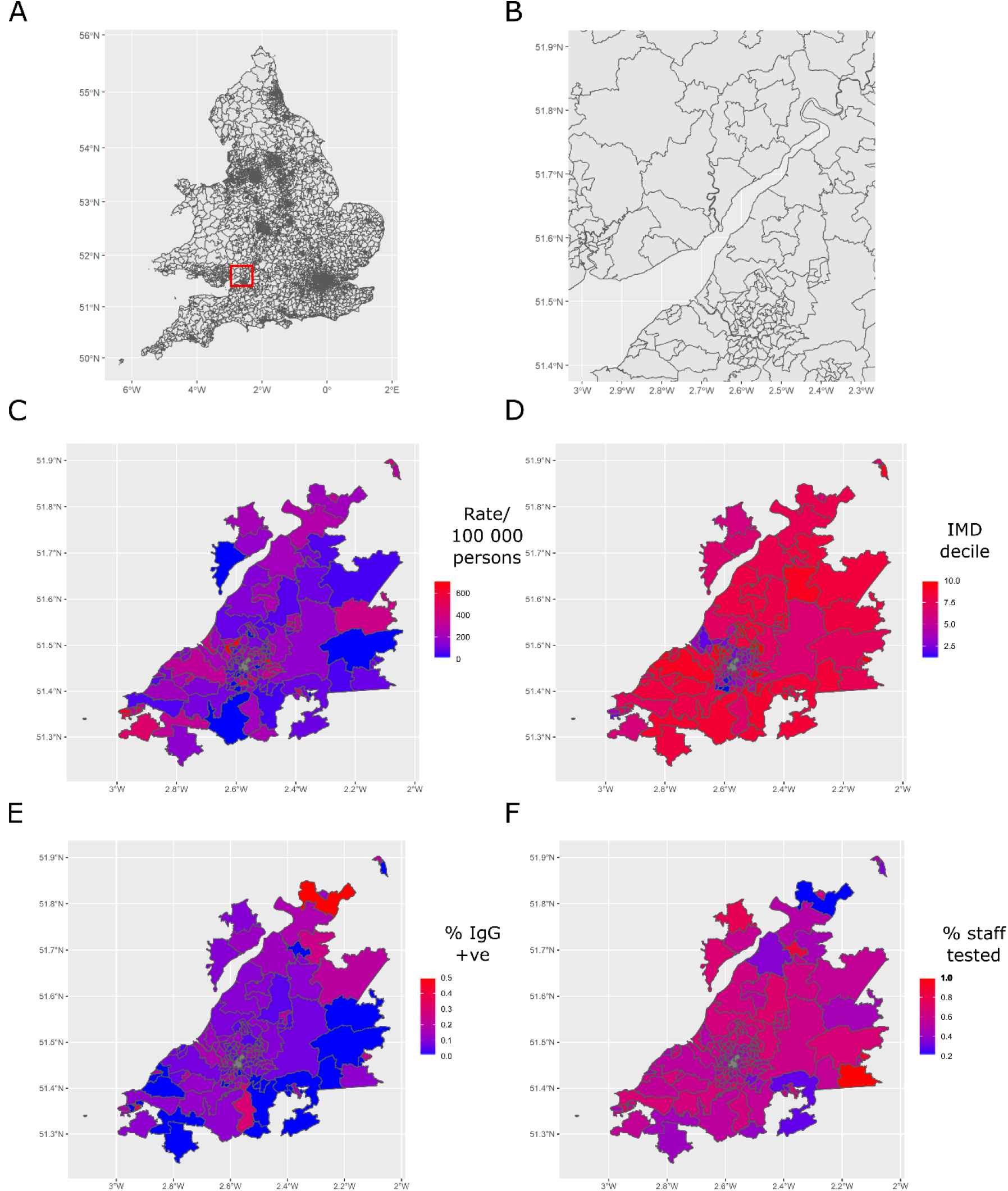
Geospatial analysis of SARS-CoV-2 IgG seroprevalence in HCWs and support staff. **(A)** Map of England and Wales highlighting the Bristol area (red square). **(B)** Bristol, South Gloucestershire, and North Somerset separated into Middle Layer Super Output Areas (MSOAs). **(C)** COVID-19 infection rate per 100,000 population across MSOA generated using data from Public Health England. **(D)** Relative deprivation across MSOA (Indices of Multiple Deprivation decile). **(E)** SARS-CoV-2 IgG seroprevalence in hospital staff in this study across MSOA. **(F)** Testing uptake by staff in this study across MSOA. **(E and F)** Data from MSOAs with <10 staff excluded.

SARS-CoV-2 seroprevalence in HCWs and support staff at the MSOA level was weakly correlated with Public Health England case rate/100 000 population (r=0.18; Figure 3).

**Figure 3:**
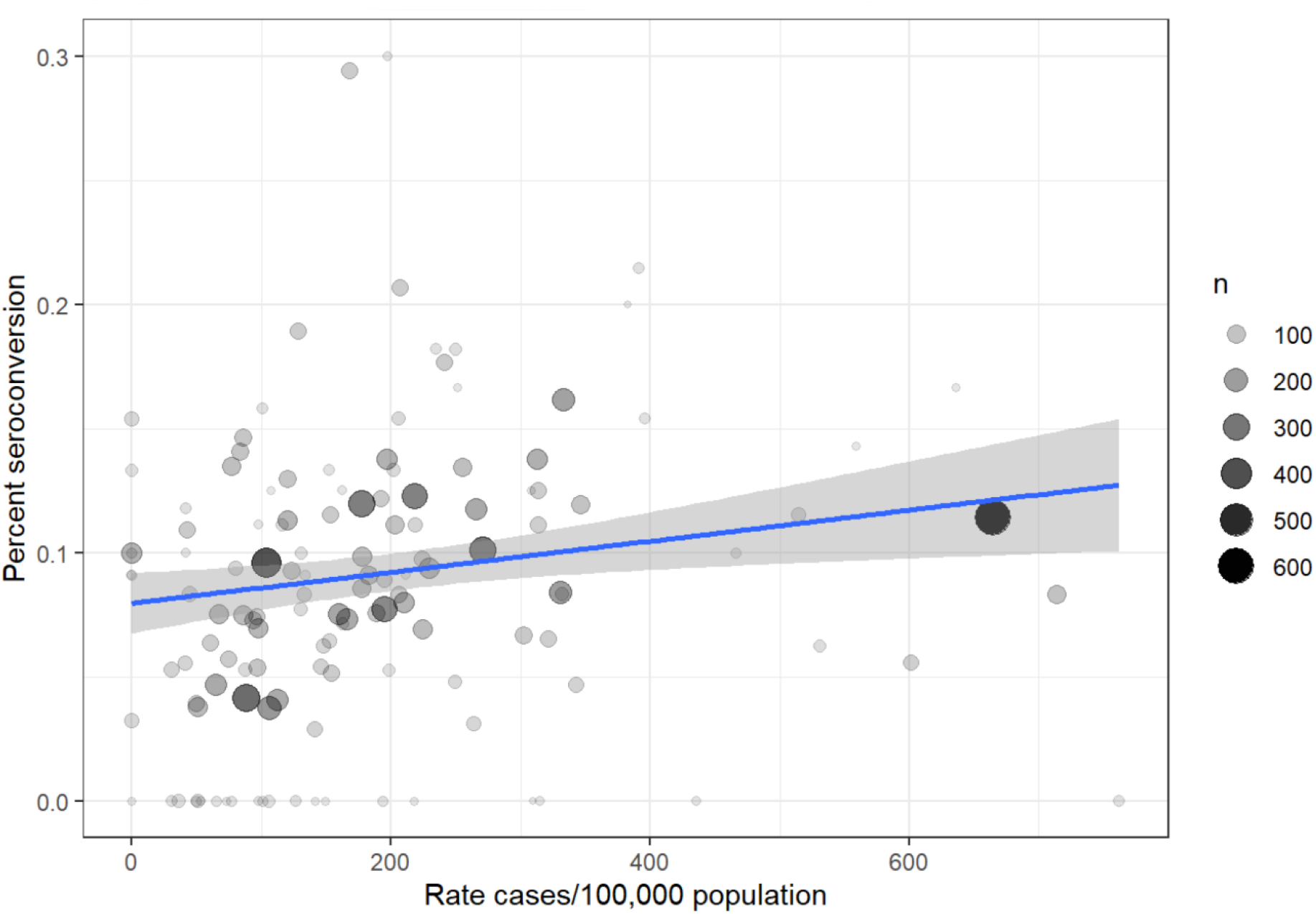
Relationship between SARS-CoV-2 IgG seroprevalence in HCWs and support staff according to middle layer super output area (MSOA) and corresponding infection rate in the general population according to PHE data. R=0.18 (Pearson’s correlation). MSOAs restricted to those with at least 10 staff member residents. Abbreviations: MSOA – Middle Layer Super Output Area; PHE – Public Health England.

### Factors associated with SARS-CoV-2 seroprevalence

Table III displays the weighted regression estimates for the assessed demographic and socioeconomic risk factors for SARS-CoV-2 seroprevalence. Black, Asian and Minority Ethnic individuals had increased odds of SARS-CoV-2 seroprevalence (adjusted OR 1.99, 95%CI: 1.69, 2.34; p<0.001) relative to White individuals. Critical care (adjusted OR 0.29, 95%CI: 0.13, 0.57; p=0.001) and theatre services (adjusted OR 0.29, 95%CI: 0.15, 0.49; p<0.001) had decreased odds of SARS-CoV-2 seroprevalence. All medicine division clusters had increased odds of seroprevalence (adjusted OR range 1.72 to 3.35; all p≤0.001). Healthcare science assistants (adjusted OR 0.35, 95%CI: 0.14, 0.73; p=0.01), healthcare science practitioners (adjusted OR 0.07, 95%CI: 0.01, 0.31; p=0.004), and specialty registrars (adjusted OR 0.62, 95%CI: 0.41, 0.91; p=0.019) had decreased odds of SARS-CoV-2 seroprevalence. Foundation year 2 doctors (adjusted OR 2.11, 95%CI: 1.40, 3.13; p<0.001), healthcare assistants (adjusted OR 1.52, 95%CI: 1.17, 1.98; p=0.002), and nurses (adjusted OR 1.35, 95%CI: 1.08, 1.69; p=0.008) had increased odds of SARS-CoV-2 seroprevalence.

**Table III:** Demographic and socioeconomic factors associated with SARS-CoV-2 IgG antibody seroprevalence in HCWs and support staff. Both unadjusted and inverse probability weight-adjusted regression data are presented. For factors with multiple categories, the 15 most populous are presented and the remaining collated into “other”, which forms the reference group. Abbreviations: IPW – inverse probability weight; OR – odds ratio; CI – confidence interval; BAME – Black, Asian and Minority Ethnic.

|  | Unadjusted model |  |  | IPW-adjusted model |  |  |
| --- | --- | --- | --- | --- | --- | --- |
| Characteristic | OR<sup>1</sup> | 95% CI<sup>1</sup> | p-value | OR<sup>1</sup> | 95% CI<sup>1</sup> | p-value |
| Ethnicity |  |  |  |  |  |  |
| White | — | — |  | — | — |  |
| BAME | 1.76 | 1.40, 2.21 | <0.001 | 1.99 | 1.69, 2.34 | <0.001 |
| Undisclosed | 1.33 | 0.76, 2.18 | 0.3 | 1.16 | 0.81, 1.61 | 0.4 |
| Gender |  |  |  |  |  |  |
| Female | — | — |  | — | — |  |
| Male | 1.01 | 0.80, 1.28 | >0.9 | 0.96 | 0.81, 1.14 | 0.7 |
| Age |  |  |  |  |  |  |
| 31-40 | — | — |  | — | — |  |
| <=20 Years | 1.06 | 0.53, 1.98 | 0.9 | 1.47 | 0.96, 2.20 | 0.071 |
| >=71 Years | 0.86 | 0.05, 4.47 | 0.9 | 0.74 | 0.17, 2.08 | 0.6 |
| 21-30 | 1.5 | 1.16, 1.95 | 0.002 | 1.64 | 1.36, 1.99 | <0.001 |
| 41-50 | 1.32 | 1.01, 1.74 | 0.045 | 1.36 | 1.11, 1.67 | 0.003 |
| 51-60 | 1.23 | 0.92, 1.64 | 0.2 | 1.45 | 1.17, 1.80 | <0.001 |
| 61-70 | 1.31 | 0.85, 1.98 | 0.2 | 1.28 | 0.94, 1.73 | 0.1 |
| Neighbourhood deprivation | 1.01 | 0.97, 1.04 | 0.7 | 0.99 | 0.96, 1.02 | 0.5 |
| Specialty |  |  |  |  |  |  |
| Other | — | — |  | — | — |  |
| Cluster 1 - Neurosurgery, Spines & Pain | 0.89 | 0.50, 1.49 | 0.7 | 0.84 | 0.51, 1.32 | 0.5 |
| Cluster 2 - Trauma & Orthopaedics | 1.46 | 0.91, 2.27 | 0.11 | 1.44 | 0.96, 2.10 | 0.067 |
| Cluster 3 | 0.96 | 0.56, 1.56 | 0.9 | 0.94 | 0.61, 1.41 | 0.8 |
| Critical Care Services | 0.31 | 0.11, 0.70 | 0.013 | 0.29 | 0.13, 0.57 | 0.001 |
| Domestics | 0.94 | 0.53, 1.65 | 0.8 | 0.99 | 0.66, 1.48 | >0.9 |
| General Surgery Services | 0.62 | 0.31, 1.12 | 0.14 | 0.62 | 0.35, 1.03 | 0.081 |
| Imaging | 0.8 | 0.46, 1.31 | 0.4 | 0.86 | 0.55, 1.28 | 0.5 |
| Maternity Services | 0.67 | 0.31, 1.31 | 0.3 | 0.75 | 0.41, 1.29 | 0.3 |
| Medicine Cluster 1 | 1.75 | 1.24, 2.43 | 0.001 | 1.72 | 1.30, 2.25 | <0.001 |
| Medicine Cluster 2 | 3.43 | 2.51, 4.67 | <0.001 | 3.35 | 2.61, 4.30 | <0.001 |
| Medicine Cluster 4 | 3.01 | 2.05, 4.37 | <0.001 | 2.84 | 2.07, 3.85 | <0.001 |
| Other Bank Services | 1.42 | 0.95, 2.07 | 0.077 | 1.17 | 0.93, 1.46 | 0.2 |
| Pathology Services | 0.51 | 0.22, 1.03 | 0.083 | 0.53 | 0.28, 0.90 | 0.028 |
| Theatre Services | 0.3 | 0.14, 0.57 | <0.001 | 0.29 | 0.15, 0.49 | <0.001 |
| Therapies Services | 1.21 | 0.72, 1.96 | 0.4 | 1.29 | 0.83, 1.93 | 0.2 |
| Role |  |  |  |  |  |  |
| Other | — | — |  | — | — |  |
| Assistant | 1.56 | 0.97, 2.44 | 0.059 | 1.39 | 0.99, 1.93 | 0.051 |
| Clerical Worker | 0.74 | 0.48, 1.12 | 0.2 | 0.81 | 0.59, 1.11 | 0.2 |
| Consultant | 0.86 | 0.52, 1.37 | 0.5 | 0.84 | 0.57, 1.23 | 0.4 |
| Foundation Year 2 | 1.46 | 0.71, 2.80 | 0.3 | 2.11 | 1.40, 3.13 | <0.001 |
| Health Care Support Worker | 2.28 | 1.27, 4.07 | 0.005 | 2.79 | 2.05, 3.82 | <0.001 |
| Healthcare Assistant | 1.57 | 1.12, 2.19 | 0.008 | 1.52 | 1.17, 1.98 | 0.002 |
| Healthcare Science Assistant | 0.41 | 0.12, 1.06 | 0.1 | 0.35 | 0.14, 0.73 | 0.01 |
| Healthcare Science Practitioner | 0.09 | 0.01, 0.45 | 0.022 | 0.07 | 0.01, 0.31 | 0.004 |
| Housekeeper | 1.67 | 0.97, 2.77 | 0.054 | 1.52 | 1.01, 2.26 | 0.041 |
| Manager | 0.89 | 0.43, 1.69 | 0.7 | 0.86 | 0.48, 1.43 | 0.6 |
| Midwife | 0.76 | 0.28, 1.94 | 0.6 | 0.59 | 0.27, 1.21 | 0.2 |
| Officer | 0.85 | 0.51, 1.36 | 0.5 | 0.84 | 0.56, 1.22 | 0.4 |
| Porter | 2.11 | 1.04, 4.00 | 0.029 | 1.57 | 1.01, 2.40 | 0.041 |
| Specialty Registrar | 0.75 | 0.43, 1.25 | 0.3 | 0.62 | 0.41, 0.91 | 0.019 |
| Staff Nurse | 1.24 | 0.94, 1.64 | 0.14 | 1.35 | 1.08, 1.69 | 0.008 |
| <sup>1</sup>OR = Odds Ratio, CI = Confidence Interval |  |  |  |  |  |  |

## Discussion

This study presents SARS-CoV-2 IgG seroprevalence data from HCWs and support staff in a low incidence setting in the UK. Data on factors linked to outcomes and risk of infection are rapidly emerging. A large UK hospital-based cohort study (ISARIC study^15^) found that older age, being male, and having chronic comorbidities such as cardiovascular disease, pulmonary disease, liver disease, kidney disease, and obesity were associated with increased mortality. An analysis of primary care records found that deprivation and either Black or South Asian ethnicity were associated with mortality, which was only partially explained by pre-existing comorbidities^16^. Emerging data suggest that, in addition to an increased risk of mortality, BAME individuals also experience higher rates of infection, which is only partially accounted for by socioeconomic and comorbid factors^17, 18^.

Studies have consistently shown higher rates of seroprevalence in HCWs. Our data from a large teaching hospital in a region that experienced lower COVID-19 disease incidence relative to the rest of the UK confirms this, although to a lesser extent than in other regions. The overall seroprevalence rate of 9.3% in the South West was lower than that reported in London (31.6%)^19^, Birmingham (24.4%)^20^, and Oxford (11%)^5^. Our data suggest that there is a weak association between staff seroprevalence and infection rate in their area of residence. Staff role, particularly roles with greater clinical contact, was associated with seroprevalence. Furthermore, we also found an association between staff seroprevalence and increased relative neighbourhood deprivation. Whilst we were unable to determine the dominant factor at play, our data highlight the complex interplay between biological, social, and economic factors that determine risk of infection during a pandemic.

As with other studies, ethnicity was associated with seroprevalence. We found a seroprevalence of 14.6% in BAME individuals, almost twice as high relative to White individuals. This observation persisted after accounting for possible bias in testing enrolment and other factors. The reason for this is likely multifactorial and could be secondary to biological, social, and/or economic factors. Interestingly, this observation was true for all staff groups apart from Medical/Dental. Our BAME Medical/Dental workforce is of equivalent seniority to their white counterparts, whereas BAME staff from other roles are often more junior. This suggests the increased seroprevalence may be related to the frequency of patient contact. Supporting this, BAME staff tended to live in more deprived neighbourhoods than their white colleagues. However, when restricting to medical and dental staff only, this difference was minimised.

As expected, working within areas of the hospital that provided care to acutely unwell patients was associated with higher rates of seroprevalence. However, in contrast to recent findings from a Danish study of HCWs^21^, seroprevalence did not associate with wards designated for COVID-19 cohorting. As observed elsewhere^20^, seroprevalence rates were low in the intensive care unit, where infection risk was likely mitigated by enhanced PPE use and probable reduced infectivity of cases that had progressed to the characterised immune-mediated disease phase. We found the highest seroprevalence rates (≥50%) in wards with known nosocomial outbreaks. Further supporting a role for transmission between staff groups, administrative and clerical staff (frequent contact with clinical staff) had higher seroprevalence rates than healthcare scientists (infrequent contact with clinical staff).

This study has limitations that must be considered when drawing conclusions. First, participants volunteered for serological testing and therefore data are subject to selection bias due to participation. To mitigate the effect of this non-random participation, inverse probability weighting was used to recover estimates that were unbiased by testing patterns on our observed characteristics. Our analysis may still be biased by other factors relating to participation that were not measured. Second, neighbourhood deprivation was analysed at the level of MSOA. The Lower Layer Super Output Area is a smaller geographical unit that offers more detailed estimates of geographical differences. However, due to sample size restrictions imposed by the testing dataset, it was not possible to perform analyses at a richer scale than MSOA. Third, it is possible that participants were more likely to attend for serological testing if they thought they had previously been infected with SARS-CoV-2. This ascertainment bias is difficult to control or adjust for in the absence of clinical data, which was not collected. Finally, HCWs are not representative of the UK population in general, nor are they representative of MSOAs, and therefore generalisability of findings is limited.

## Conclusion

This study presents some of the first SARS-CoV-2 IgG seroprevalence data in HCWs and support staff in a low incidence setting in the UK. It provides further evidence that BAME individuals are at increased risk of infection with SARS-CoV-2 and this may be influenced by work role and domestic risk factors. Transmission between staff groups in the first part of the pandemic is clear – both in the high incidence in ward outbreaks and higher rates in those non-clinical staff working directly with clinical staff. Using weighted models, medical subspecialities, junior medical staff, nursing staff and estate staff were at the highest risk. Identifying HCWs at increased risk of infection with SARS-CoV-2 will support the implementation of targeted interventions designed to ensure the entire workforce is protected during future community and hospital onset COVID-19 outbreaks. As hospitals consider routine staff PCR testing for SARS-CoV-2 they should ensure that they account for the decreased uptake in certain staff groups and ensure equity as much as possible. Multi-site longitudinal studies of HCWs are needed to further clarify the demographic and socioeconomic characteristics that have the biggest influence on risk of infection and COVID-19 disease.

## Data Availability

Data available within the article or its supplementary materials.

## Acknowledgements

We are grateful to North Bristol NHS Trust staff for their participation and providing data. We are grateful to Benjamin Pope for his technical support during this project.

## Conflict of interest

None declared.

## Funding

This research did not receive any specific grant from funding agencies in the public, commercial, or not-for-profit sectors.

## Notes

### Competing Interest Statement

The authors have declared no competing interest.

## References

1. Zhu N, Zhang D, Wang W et al. A Novel Coronavirus from Patients with Pneumonia in China, 2019. N Engl J Med 2020; 382: 727–33.

2. Covid-19 dashboard by the Center for Systems Science and Engineering (CSSE) at Johns Hopkins University (JHU). Available at https://coronavirus.jhu.edu/map.html. [Accessed on 30th October 2020]. 2020.

3. Carter B, Collins JT, Barlow-Pay F et al. Nosocomial COVID-19 infection: examining the risk of mortality. The COPE-Nosocomial Study (COVID in Older PEople). J Hosp Infect 2020; 106: 376–84.

4. Burki T England and Wales see 20?000 excess deaths in care homes. Lancet 2020; 395: 1602-.

5. Eyre DW, Lumley SF, O’Donnell D et al. Differential occupational risks to healthcare workers from SARS-CoV-2 observed during a prospective observational study. Elife 2020; 9: e60675.

6. Liu M, Cheng S-Z, Xu K-W et al. Use of personal protective equipment against coronavirus disease 2019 by healthcare professionals in Wuhan, China: cross sectional study. BMJ 2020; 369: m2195–m.

7. Nguyen LH, Drew DA, Graham MS et al. Risk of COVID-19 among front-line health-care workers and the general community: a prospective cohort study. Lancet Public Health 2020; 5: e475–e83.

8. Steel N, Ford JA, Newton JN et al. Changes in health in the countries of the UK and 150 English Local Authority areas 1990-2016: a systematic analysis for the Global Burden of Disease Study 2016. Lancet 2018; 392: 1647–61.

9. Deaths involving COVID-19 by local area and socioeconomic deprivation: deaths occurring between 1 March and 30 June 2020. Office for National Statistics 2020.

10. Griffith G, Morris TT, Tudball M et al. Collider bias undermines our understanding of COVID-19 disease risk and severity. medRxiv 2020; doi 10.1101/2020.05.04.20090506: 2020.05.04.20090506.

11. Mansournia MA, Altman DG Inverse probability weighting. BMJ 2016; 352: i189.

12. McLennan D NS, Noble M, Plunkett E, Wright G, Gutacker, N. The English Indices of Deprivation: Technical Report. Ministry of Housing, Communities, and Local Government 2019.

13. English indices of deprivation 2019. Available at https://www.gov.uk/government/statistics/english-indices-of-deprivation-2019. [Accessed on 5th October 2020]. Ministry of Housing, Communities and Local Government.

14. PHE. Coronavirus (COVID-19) in the UK. Available at https://coronavirus.data.gov.uk/. [Accessed on 5th October 2020]. Public Health England 2020.

15. Docherty AB, Harrison EM, Green CA et al. Features of 20?133 UK patients in hospital with covid-19 using the ISARIC WHO Clinical Characterisation Protocol: prospective observational cohort study. BMJ 2020; 369: m1985–m.

16. Williamson EJ, Walker AJ, Bhaskaran K et al. Factors associated with COVID-19-related death using OpenSAFELY. Nature 2020; 584: 430–6.

17. de Lusignan S, Dorward J, Correa A et al. Risk factors for SARS-CoV-2 among patients in the Oxford Royal College of General Practitioners Research and Surveillance Centre primary care network: a cross-sectional study. Lancet Infect Dis 2020; 20: 1034–42.

18. Niedzwiedz CL, O’Donnell CA, Jani BD et al. Ethnic and socioeconomic differences in SARS-CoV-2 infection: prospective cohort study using UK Biobank. BMC Med 2020; 18: 160-.

19. Grant JJ, Wilmore SMS, McCann NS et al. Seroprevalence of SARS-CoV-2 antibodies in healthcare workers at a London NHS Trust. Infect Control Hosp Epidemiol 2020; doi 10.1017/ice.2020.402: 1-3.

20. Shields A, Faustini SE, Perez-Toledo M et al. SARS-CoV-2 seroprevalence and asymptomatic viral carriage in healthcare workers: a cross-sectional study. Thorax 2020; doi 10.1136/thoraxjnl-2020-215414: thoraxjnl-2020-215414.

21. Iversen K, Bundgaard H, Hasselbalch RB et al. Risk of COVID-19 in health-care workers in Denmark: an observational cohort study. Lancet Infect Dis 2020; doi 10.1016/S1473-3099(20)30589-2:S1473-3099(20)30589-2.

