## Supplementary Data for "Seroprevalence of SARS-CoV-2 IgG in healthcare workers and other staff at North Bristol NHS Trust: a sociodemographic analysis"

### Supplementary material

**Supplementary table I: Odds of attending for testing according to demographic and socioeconomic factors.** These data were used to generate the inverse probability weighted model for assessing factors associated with SARS-CoV-2 seroprevalence. For factors with multiple categories, the 15 most populous are presented and the remaining collated into "other", which forms the reference group.

| Characteristic | OR[1] | 95% CI[1] | p-value |
|---|---|---|---|
| Ethnicity | | | |
| White | — | — | |
| BAME | 0.85 | 0.76, 0.96 | 0.007 |
| Undisclosed | 0.51 | 0.41, 0.62 | <0.001 |
| Gender | | | |
| Female | — | — | |
| Male | 0.82 | 0.74, 0.91 | <0.001 |
| Age | | | |
| 31-40 | — | — | |
| <=20 Years | 1.04 | 0.77, 1.41 | 0.8 |
| >=71 Years | 0.55 | 0.28, 1.02 | 0.064 |
| 21-30 | 1.15 | 1.03, 1.29 | 0.016 |
| 41-50 | 1.33 | 1.17, 1.50 | <0.001 |
| 51-60 | 1.26 | 1.11, 1.43 | <0.001 |
| 61-70 | 0.94 | 0.79, 1.13 | 0.5 |
| IMD | 1 | 0.99, 1.02 | 0.6 |
| Specialty | | | |
| Other | — | — | |
| Cluster 1 - Neurosurgery, Spines & Pain | 1.83 | 1.39, 2.44 | <0.001 |
| Cluster 2 - Trauma & Orthopaedics | 1.93 | 1.47, 2.56 | <0.001 |
| Cluster 3 | 1.32 | 1.04, 1.68 | 0.025 |
| Critical Care Services | 0.96 | 0.72, 1.27 | 0.8 |
| FOH Domestics | 0.74 | 0.57, 0.96 | 0.024 |
| General Surgery Services | 1.86 | 1.41, 2.48 | <0.001 |
| Imaging | 1.43 | 1.14, 1.82 | 0.002 |
| Maternity Services | 2.03 | 1.46, 2.88 | <0.001 |
| Medicine Cluster 1 | 1.46 | 1.21, 1.77 | <0.001 |
| Medicine Cluster 2 | 1.24 | 1.01, 1.52 | 0.04 |
| Medicine Cluster 4 | 1.25 | 0.98, 1.60 | 0.076 |
| Other Bank Services | 0.17 | 0.14, 0.19 | <0.001 |
| Pathology Services | 0.76 | 0.62, 0.93 | 0.007 |
| Theatre Services | 1.53 | 1.22, 1.93 | <0.001 |
| Therapies Services | 1.74 | 1.33, 2.29 | <0.001 |
| Role | | | |
| Other | — | — | |
| Assistant | 0.84 | 0.68, 1.05 | 0.13 |
| Clerical Worker | 0.86 | 0.72, 1.01 | 0.066 |
| Consultant | 1.12 | 0.91, 1.39 | 0.3 |
| Foundation Year 2 | 0.97 | 0.70, 1.32 | 0.8 |
| Health Care Support Worker | 0.79 | 0.61, 1.01 | 0.063 |
| Healthcare Assistant | 0.69 | 0.58, 0.82 | <0.001 |
| Healthcare Science Assistant | 0.91 | 0.68, 1.22 | 0.5 |
| Healthcare Science Practitioner | 0.96 | 0.73, 1.28 | 0.8 |
| Housekeeper | 0.75 | 0.57, 0.98 | 0.033 |
| Manager | 0.98 | 0.75, 1.30 | 0.9 |
| Midwife | 0.62 | 0.41, 0.91 | 0.018 |
| Officer | 0.83 | 0.68, 1.02 | 0.074 |
| Porter | 0.6 | 0.43, 0.82 | 0.001 |
| Specialty Registrar | 0.59 | 0.48, 0.72 | <0.001 |
| Staff Nurse | 1.14 | 0.99, 1.31 | 0.078 |

1 OR = Odds Ratio, CI = Confidence Interval

**Supplementary table II: Seniority of Medical and Dental HCWs stratified by ethnicity.** Data are restricted to those staff members that comprise the Medical and Dental group.

| Characteristic | BAME N = 309 | | Undisclosed N = 126 | | White N = 1,313 | |
|---|---|---|---|---|---|---|
| Role | n | % | n | % | n | % |
| Consultant | 119 | 39% | 46 | 37% | 414 | 32% |
| Specialty Registrar | 116 | 38% | 48 | 38% | 520 | 40% |
| Specialty Doctor | 19 | 6.1% | 2 | 1.6% | 30 | 2.3% |
| Trust Grade Doctor | 2 | 0.6% | 1 | 0.8% | 8 | 0.6% |
| Foundation Year 2 | 39 | 13% | 24 | 19% | 247 | 19% |
| Foundation Year 1 | 12 | 3.9% | 5 | 4% | 83 | 6.3% |
| Statistics presented: n (%) | | | | | | |

**Supplementary table III: SARS-CoV-2 IgG seroprevalence in HCWs and support staff from inpatient wards and dialysis units managed by the Trust**. Data represent proportion of the tested population. Specialty of ward area reported in place of ward names.

| Variable | Serology +ve | | Total |
|---|---|---|---|
| | n | % | |
| Dialysis Unit 1 | 10 | 58.8% | 17 |
| Cardiology | 22 | 52.4% | 42 |
| Elderly Care step-down | 20 | 50.0% | 40 |
| Elderly Care | 19 | 38.8% | 49 |
| Elderly Care | 13 | 31.0% | 42 |
| Dialysis Unit 2 | 6 | 30.0% | 20 |
| General Medicine | 12 | 27.9% | 43 |
| Elderly Care step-down | 10 | 27.8% | 36 |
| Urology | 8 | 25.8% | 31 |
| Neurorehabilitation Unit | 1 | 25.0% | 4 |
| Surgery | 9 | 24.3% | 37 |
| Stroke | 12 | 23.1% | 52 |
| Surgery/Orthopaedics | 8 | 22.2% | 36 |
| Complex Medical + COVID-19 | 9 | 20.9% | 43 |
| Gastroenterology | 9 | 18.8% | 48 |
| Acute Medical Unit | 18 | 16.2% | 111 |
| Orthopaedics | 6 | 15.4% | 39 |
| Respiratory + COVID-19 | 6 | 13.6% | 44 |
| Surgery | 7 | 13.0% | 54 |
| General Medicine | 5 | 11.1% | 45 |
| Emergency Dept | 4 | 10.8% | 37 |
| Surgery | 5 | 8.8% | 57 |
| Neurosurgery | 4 | 8.7% | 46 |
| Orthopaedics | 3 | 7.9% | 38 |
| Dialysis Unit 3 | 1 | 7.1% | 14 |
| Emergency Dept – Nursing | 12 | 6.9% | 173 |
| Vascular | 3 | 6.7% | 45 |
| Neurology | 2 | 5.0% | 40 |
| Renal | 2 | 5.0% | 40 |
| Dialysis Unit 4 | 1 | 4.8% | 21 |
| Surgical | 1 | 3.4% | 29 |
| Adult ICU | 4 | 2.5% | 159 |
| Neonatal ICU | 2 | 2.2% | 93 |
| Women's Health | 0 | 0.0% | 24 |
| Dialysis Unit 5 | 0 | 0.0% | 14 |
